# Demographic, temporal, and spatial analysis of human rabid animal bite cases in Mymensingh District, Bangladesh

**DOI:** 10.1101/2024.05.12.24307159

**Authors:** Chandra Shaker Chouhan, Abu Raihan, Md. Manik Mia, Subarna Banerjee, Ishmam Shahriar, Proggananda Nath, Jasim M. Uddin, Md. Amimul Ehsan, Michael P. Ward, A. K. M. Anisur Rahman

## Abstract

**Objective:** This study aimed to analyze the demographic, temporal, and spatial characteristics of rabid animal bite (RAB) cases in humans across 12 upazilas within Mymensingh district of Bangladesh.

**Methods:** Retrospective hospital-based data from RAB cases for 2022 and 2023 were collected from S.K Hospital. The dataset included information on victim demographics, bite details, vaccination information, and Rabies Immune Globulin (RIG) administration. Additionally, monthly case counts from 2016 to 2023 were sourced and analyzed to identify trends. Descriptive statistics and time series analysis using the seasonal decomposition technique were conducted. The risk maps for rabid animal bites in 2022 and 2023 were generated using a standardized incidence rate ratio (SIRR) approach.

**Findings:** An almost two-fold increase in the proportion of category 3 bites receiving Rabies Immune Globulin (RIG) from 3.6% in 2022 to 6.5% in 2023 was noted. Only 9.7% of bite cases in 2022 and 16.9% in 2023 received the vaccine promptly after the incident. However, the majority received vaccines within the first 24 hours after being bitten. Moreover, significant seasonal patterns and year-wise increasing trends in RAB cases were observed. Males and individuals <10 years old had a higher risk of being bitten. Dogs (48.2% in 2022) and cats (52.6% in 2023) were identified as the primary animals responsible for the bites. Notably, the legs were the body part most frequently bitten. The bites risk map identified four high risk upazilas.

**Conclusion:** There is a significant gap in ensuring timely vaccination delivery. Study results also suggest other potential improvements in healthcare practices or treatment protocols. Increasing RAB cases highlights the need to enhance surveillance and control measures. Targeted awareness campaigns and preventive measures tailored to high-risk groups − including males, children <10 years old, dogs and cats − are imperative. Coordinated efforts among healthcare professionals, policymakers, and community stakeholders are crucial to effectively mitigate the incidence of RAB cases, safeguarding public health and eradicate dog mediated rabies by 2030 in the region.

## 1. Introduction

Rabies, caused by Lyssavirus type 1 of the Rhabdoviridae family, is a fatal viral infection of the central nervous system that results in progressive encephalomyelitis (1–3). The virus is typically present in the saliva of clinically ill mammals, which can then be transmitted to other mammals (including humans) through a bite, scratch, or lick (4). Rabies manifests in two epidemiological conditions: urban, primarily transmitted by dogs or less commonly by cats; and sylvatic, carried by wolves, foxes, raccoons, weasels, and bats (5).

In humans, transmission mainly occurs from canines, as well as from cats, mongoose, bats, and in rare cases, farm animals (6). Globally, > over 1.4 billion people are at risk of rabies, and approximately 45% of rabies-related deaths occur in Asia (7). Human rabies cases that result from dog bites constitute 97% of all cases (8). Bangladesh ranks third after India and China in terms of rabies mortalities, with >2100 people dying annually (9). Dogs (90%), cats (6%), jackals (3%), and mongoose (1%) were reported to be responsible for human bites in Bangladesh (9).

The most effective strategy for preventing human rabies is to target the animal reservoir, using mass dog vaccination to stop transmission (10). The World Health Organization (WHO) and its partners, including the World Organization for Animal Health (OIE), the Food and Agriculture Organization of the United Nations (FAO) and the Global Alliance for Rabies Control (GARC), have adopted a goal to eliminate dog-mediated human rabies by 2030 by controlling the disease in dogs (11). In developed countries, concerted efforts such as mass vaccination of dogs and administering oral vaccines to wildlife have significantly contributed to the eradication of rabies among dogs, resulting in a subsequent decrease in human rabies cases (12). Nevertheless, timely wound management is crucial as an emergency medical response in disease control measures (13).

Rabies cases often do not fully represent the disease burden due to negligence and possible underreporting in specific areas (14). Unreported cases and a lack of understanding of epidemiological trends pose a significant threat to the efficient execution of preventive and control measures (15, 16). In Bangladesh, inadequate surveillance and reporting systems, together with widespread misconceptions among the general population, hinder proper documentation of epidemiological data on animal bite cases and fatalities (17). Furthermore, limited financial resources impede the conduct of surveys to determine rabies incidence in animals, and there are limited opportunities for post-treatment studies of patients. Additionally, the lack of laboratory confirmation in the identification of rabid animals, primarily due to financial and logistical limitations, further exacerbates the issues (18).

Implementing measures to decrease animal bite incidence is one of the critical approaches to controlling rabies (2). A comprehensive strategy for eradicating rabies in Bangladesh has been in place since June 2010 (19). Since 2011, these initiatives have resulted in a declining pattern in human rabies-related fatalities in recent years. However, the number of cases remains notable (15). Understanding the demographic, spatial and temporal patterns of rabies in humans is crucial for assessing risks and developing targeted interventions. To do so, the current study investigated the demographic, temporal, and spatial characteristics of human rabid animal bite (RAB) cases in the Mymensingh district.

## 2. Methodology

### 2.1. Study design and data source

A retrospective hospital-based cross-sectional study was conducted to collect secondary data on cases of animal bites that occurred from January 2022 to December 2023. The data were retrieved from registers maintained at the Government S.K. Hospital in Mymensingh District. The data included victim’s address, age, sex, location of the bite, type of animal involved, date of the attack, date of vaccination, and details on the amount of HRIG provided, as well as the injury category. Additionally, we collected monthly total number of cases from 2016 to 2023 to assess the seasonal patterns and yearly trends in animal bite cases. No data were available for the year 2020 due to the COVID-19 pandemic.

### 2.2. Data processing and descriptive analysis

The data for each bite case were initially entered into Microsoft Excel 2017. Subsequently, the data were imported into Jupyter Notebook, Python, for further analysis. Descriptive statistics for continuous variables, such as age and the amount of Rabies Immune Globulin (RIG) administered, were generated using the Pandas library’s ‘description ()’ command in Python. For categorical variables, frequency distributions were created using the Pandas library’s ‘value counts ()’ command. The proportion of each category in categorical variables was calculated by using the Pandas library’s ‘value_counts ()’ command with the ‘normalize=True’ parameter.

### 2.3. Temporal distribution analysis

We used a time series analysis to explore temporal patterns and trends in the dataset. The first step involved preparing the time series data, which consisted of monthly counts of rabid animal bite cases. The data were loaded as a pandas Data Frame, and the index was set to a date time format to facilitate time-based analyses. An exploratory data analysis was then conducted to gain insights into the general characteristics of the time series. This included visualizations such as line plots and histograms to understand the distribution of cases over time. To identify underlying patterns within the time series, a seasonal decomposition using an additive model was performed via the ’seasonal_decompose’ function from the ’statsmodels’ library. This facilitated the extraction of temporal components including trend, seasonality, and residuals. To assess the stationarity of the time series, the Augmented Dickey-Fuller test was employed. This test helps determine if the time series exhibits stable behaviour over time or requires differencing.

### 2.4. RAB risk mapping analysis

A standardized risk ratio map was created using the following steps in Python:

#### i. Global incidence calculation

The global incidence of rabid bites, representing the total number of cases divided by the total population across all Upazilas, was estimated using the following formula:

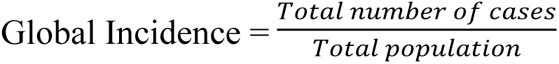

#### ii. Expected number of cases for each Upazila

The expected number of cases for each Upazila was calculated by multiplying the global incidence and the population of each Upazila as shown in equation 1:

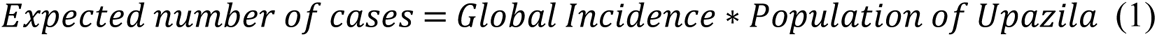

#### iii. Standardized incidence rate ratio (SIRR) calculation

The Standardized Incidence Rate Ratio (SIRR) was then calculated for each Upazila by dividing the reported number of RAB cases by the expected number of cases in each Upazila according to previously described method (20) using equation (2).

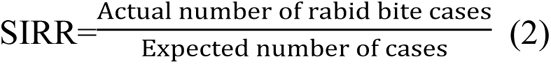

#### iv. Geospatial data loading

The Bangladesh Upazila level shape file was downloaded from the GADM maps and data (https://gadm.org/). The Geopandas library was used to read a shape file containing the geographic boundaries of Upazilas in Bangladesh.

#### v. Upazila selection and filtering & geospatial data merging

The Upazilas of Mymensingh district were selected and filtered to visualize the Map of Mymensingh district. The filtered geospatial data (Upazila boundaries) and the data containing SIRR values were merged based on the common Upazila names.

#### vi. Risk ratio map creation & annotation, and visualization

Using ’matplotlib’ and ’geopandas’, a choropleth map was created where the fill colour represented the SIRR values of each Upazila. The map was visualized with distinct colours indicating variations in standardized risk ratios. Upazila names were annotated at the centroids of their respective boundaries to enhance map readability.

## 3. Results

### 3.1. Descriptive statistics

The S. K. Hospital in the Mymensingh district documented 14,943 cases of human rabid animal bites in 2022 and 18,998 cases in 2023. However, only 9.7% in 2022 and 16.9% in 2023 received the vaccine promptly after the incident. The majority received the vaccine within the first 24 hours after being bitten **(Table 1)**

**Table 1.** Time gap between animal bite and anti-rabies vaccination.

| Year | Day 0 | Day 1 | Day 2 | Day 3 | Day 4 | Day 5 | ≥ Day 6 |
| --- | --- | --- | --- | --- | --- | --- | --- |
| 2022 | 1449 | 9987 | 2458 | 628 | 238 | 101 | 82 |
|  | (9.7%) | (66.8%) | (16.5%) | (4.2%) | (1.6%) | (0.7%) | (0.5%) |
| CP | 9.7%) | 76.5% | 93.0 % | 97.2 % | 98.8% | 99.5% | 100% |
| 2023 | 3209<br>(16.9%) | 12859<br>(67.7%) | 1994<br>(10.5%) | 588<br>(3.1%) | 170<br>(0.9%) | 104<br>(0.5%) | 74<br>(0.4%) |
| CP | 16.9% | 84.6% | 95.1% | 98.2% | 99.1% | 99.6% | 100% |

CP: Cumulative proportion

In 2022, the rabies vaccine was administered in four doses, scheduled at days 0, 3, 7, and 28. However, in 2023 the vaccination protocol was revised, with only three doses given on days 0, 3, and 21. Despite the change, almost all individuals completed the second dose of the vaccine course in both 2022 and 2023. The proportion of people who did not complete the entire vaccination course decreased from 10.3% in 2022 to 7.8% in 2023 **(Table 2).**

**Table 2.** Completion status of full vaccination regimen.

| Year | Day 0 |  | Day 3 |  | Day 7 |  | Day 21 |  | Day 28 |  |
| --- | --- | --- | --- | --- | --- | --- | --- | --- | --- | --- |
|  | Yes | No | Yes | No | Yes | No | Yes | No | Yes | No |
| 2022 | 14943<br>(100%) | 0 | 14906<br>(99.7%) | 37<br>(0.3%) | 14173<br>(94.9%) | 770<br>(5.2%) | — | — | 13398<br>(89.7%) | 1545<br>(10.3%) |
| 2023 | 18998<br>(100%) | 0 | 18958<br>(99.8%) | 40<br>(0.2%) | — | — | 17510<br>(92.2%) | 1488<br>(7.8%) | — | — |

The demographic characteristics of rabid animal bite cases are presented in **Table 3**. In 2022, there were 490 cases (3.6%) categorized as type 3 bites, which increased to 6.5% in 2023, all of which exclusively received Rabies Immune Globulin (RIG). The study found that the average age of the cases was 25.4 years, with an interquartile range 10 to 37.0 years, thus we classify age into four categories: up to 10 years, 11-25 years, 26-37 years, and above 37 years. In both 2022 and 2023 the highest percentage of bite cases was in the age group <10 years old, 27.4% and 30.1%, respectively. More cases were observed in males, making up 63.2% of bite cases in 2022 and 61.2% in 2023. In 2022, dogs were the most common animals involved in the bites, accounting for 48.2%, whereas in 2023 cats were the majority, making up 52.6%. Besides dogs and cats, foxes, mongoose, and monkeys were also involved in bite cases. The most common biting site was the right leg (31.5% in 2022 and 30.9% in 2023), followed by the left leg (28.5% in 2022 and 25.4% in 2023). Additionally, there were reports of bites to the face and even the eye.

**Table 3.** Demographic distribution of rabid animal bite cases reported in Mymensingh district, Bangladesh.

| Variables | Category level | Number of total cases<br>(33,941) |  |
| --- | --- | --- | --- |
|  |  | Year 2022<br>(14,943) | Year 2023<br>(18,998) |
| Injury category | Category 2 | 14405 (96.4%) | 17767 (93.5%) |
|  | Category 3 | 538 (3.6%) | 1231 (6.48%) |
| Age (Years) | Up to 10 | 4091 (27.4%) | 5710 (30.1%) |
|  | 11-25 | 3709 (24.8%) | 4677 (24.6%) |
|  | 26-37 | 3416 (22.9%) | 4222 (22.2%) |
|  | Above 37 | 3727 (24.94%) | 4389 (23.1%) |
| Sex | Male | 9448 (63.2%) | 11627 (61.2%) |
|  | Female | 5495 (36.8%) | 7371 (38.8%) |
| Animals involved | Dog | 7196 (48.2%) | 8132 (42.8%) |
|  | Cat | 7107 (47.6%) | 9983 (52.6%) |
|  | Fox | 404 (2.7%) | 560 (3.0%) |
|  | Monkey | 193 (1.3%) | 255 (1.3%) |
|  | Mongoose | 43 (0.3%) | 68 (0.4%) |
| Biting sites | Right Leg | 4708 (31.5%) | 5863 (30.9%) |
|  | Left Leg | 4255 (28.5%) | 4818 (25.4%) |
|  | Right Hand | 2467 (16.5%) | 3406 (17.9%) |
|  | Left Hand | 2243 (15.0%) | 3176 (16.7%) |
|  | Thigh | 281 (1.9%) | 429 (2.3%) |
|  | Back | 262 (1.8%) | 376 (2.0%) |
|  | Hip | 245 (1.6%) | 266 (1.4%) |
|  | Face | 182 (1.2%) | 253 (1.3%) |
|  | Abdomen | 81 (0.54%) | 93 (0.49%) |
|  | Both hand | 61 (0.41%) | 72 (0.38%) |
|  | Both Leg | 52 (0.34%) | 59 (0.31%) |
|  | Head | 49 (0.33%) | 61 (0.32%) |
|  | Neck | 29 (0.19%) | 85 (0.45%) |
|  | Thorax | 18 (0.12%) | 21 (0.11%) |
|  | Eye | 11 (0.07%) | 19 (0.1%) |

### 3.2. Month-wise distribution of rabid animal bite cases

The monthly distribution of rabid animal bite cases is shown in **Figure 1**. The lowest number of cases was observed in June in both 2022 (6.9%) and 2023 (6.5%), while the highest number of cases was reported in November in both 2022 (11.2%) and 2023 (11.4%).

**Figure 1:**
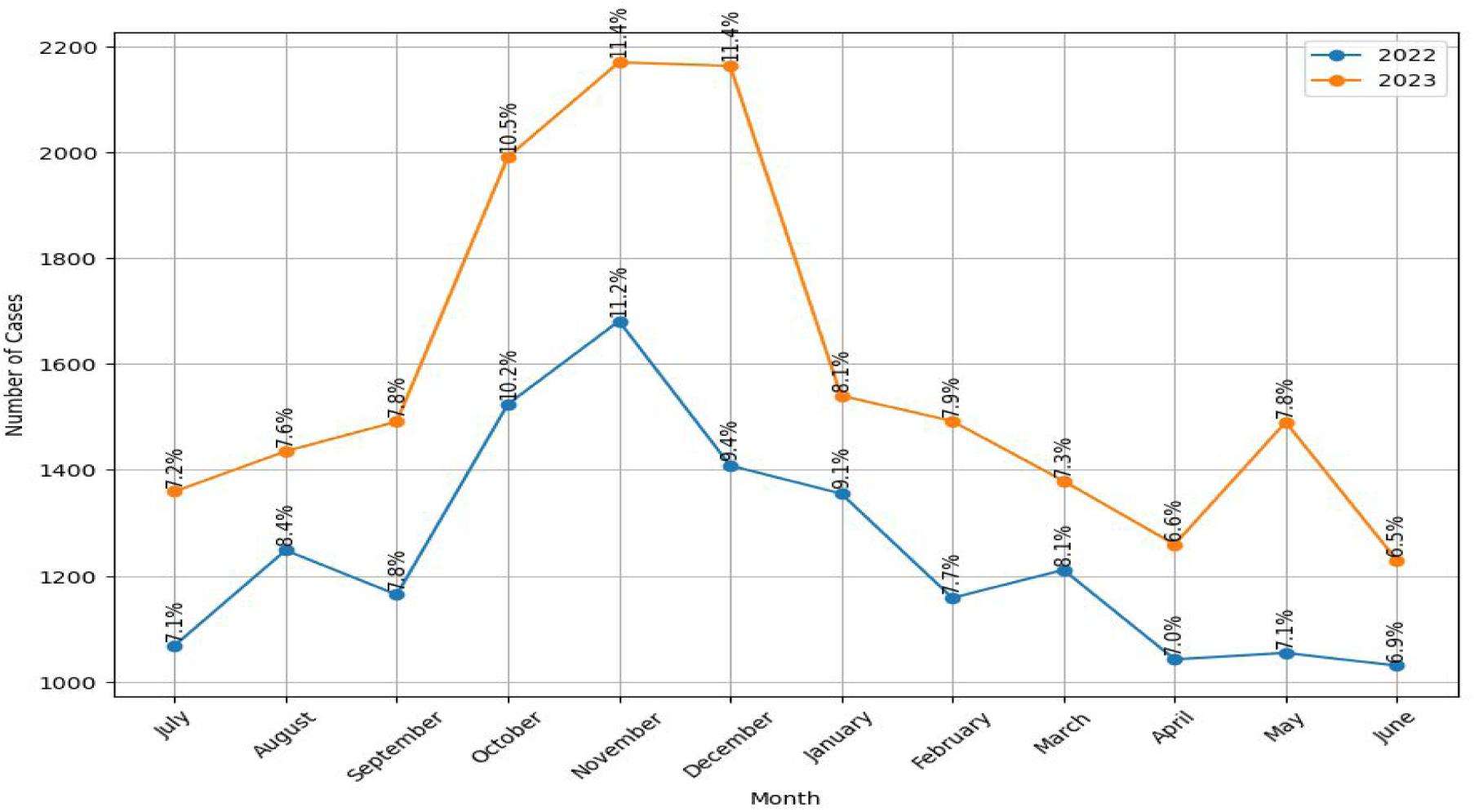
Month wise distribution of rabid animal bite cases in 2022 and 2023

### 3.3. Spatial distribution of animal bite cases

Rabid animal bite cases were reported from 12 Upazilas in the Mymensingh district. The highest number of cases was recorded in Mymensingh Sadar Upazila, accounting for 56.5% (8,441 cases) in 2022 and 52.3% (9,930 cases) in 2023. Conversely, the lowest number of cases was reported in Dhobaura Upazila with 0.70% (105 cases) in 2022 and in Gaffargaon with 0.50% (95 cases) in 2023. Notably, during 2022 and 2023, only four upazilas (Mymensingh Sadar, Phulpur, Trishal, and Muktagaccha) had a SIRR >1. Furthermore, only two upazilas (Mymensingh Sadar and Trishal) exhibited a SIRR >1 in both years **(Table 4).**

**Table 4.**
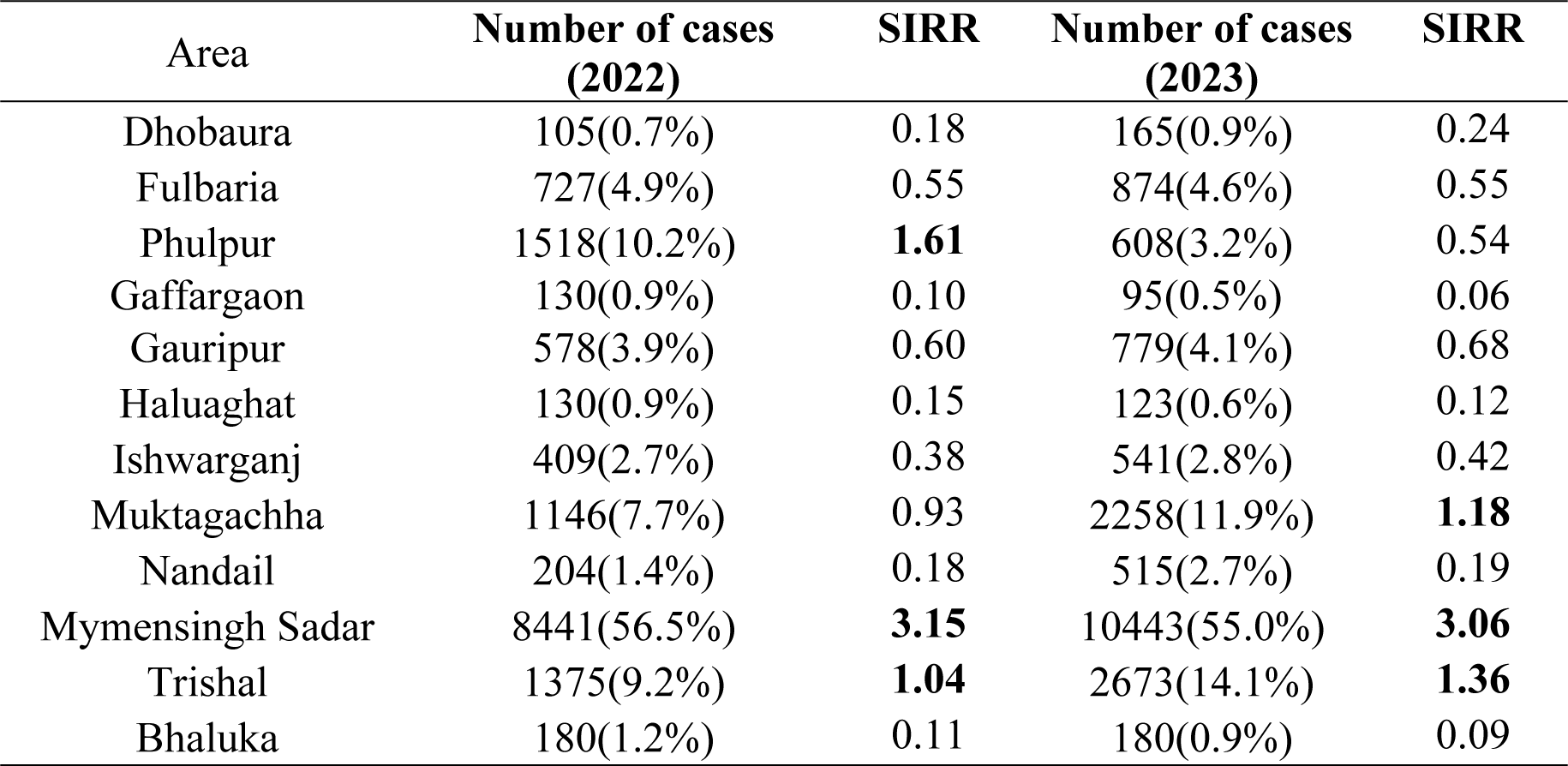
Spatial distribution of rabid animal bite cases in humans in the Mymensingh district during 2022-2023.

| Area | Number of cases<br>(2022) | SIRR | Number of cases<br>(2023) | SIRR |
| --- | --- | --- | --- | --- |
| Dhobaura | 105(0.7%) | 0.18 | 165(0.9%) | 0.24 |
| Fulbaria | 727(4.9%) | 0.55 | 874(4.6%) | 0.55 |
| Phulpur | 1518(10.2%) | **1.61** | 608(3.2%) | 0.54 |
| Gaffargaon | 130(0.9%) | 0.10 | 95(0.5%) | 0.06 |
| Gauripur | 578(3.9%) | 0.60 | 779(4.1%) | 0.68 |
| Haluaghat | 130(0.9%) | 0.15 | 123(0.6%) | 0.12 |
| Ishwarganj | 409(2.7%) | 0.38 | 541(2.8%) | 0.42 |
| Muktagachha | 1146(7.7%) | 0.93 | 2258(11.9%) | **1.18** |
| Nandail | 204(1.4%) | 0.18 | 515(2.7%) | 0.19 |
| Mymensingh Sadar | 8441(56.5%) | **3.15** | 10443(55.0%) | **3.06** |
| Trishal | 1375(9.2%) | **1.04** | 2673(14.1%) | **1.36** |
| Bhaluka | 180(1.2%) | 0.11 | 180(0.9%) | 0.09 |

### 3.4. Rabid animal bite risk mapping

The highest risk of rabid animal bites was observed in 2022 at Mymensingh Sadar Upazila (IRR: 3.15), followed by Phulpur (IRR:1.61) and Trishal (IRR: 1.04). In 2023, the highest risk was also in Mymensingh Sadar (IRR: 3.06), followed by Muktagaccha (IRR: 1.18) and Trishal (IRR: 1.36). The risk map of rabid animal bite cases is presented in **Figure 2(a-b).**

**Figure 2 (a-b):**
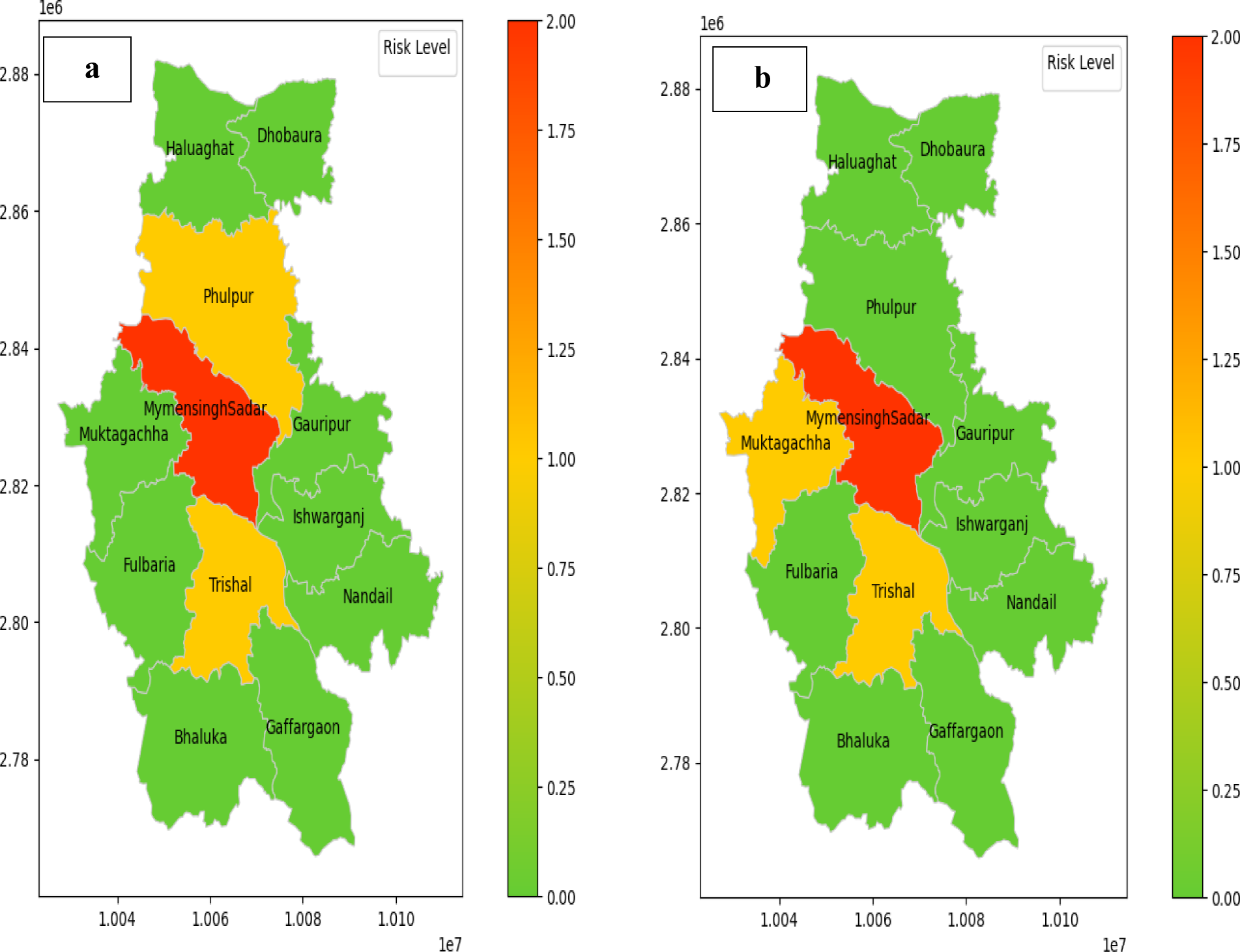
Rabid animal bite risk map during 2022 (a) and 2023 (b) in different upazilas of Mymensingh district

### 3.5. Seasonality and annual trend in rabid animal bite cases from 2016 to 2023

The time series analysis indicates both seasonal variation and an increasing trend in rabid animal bite cases **(Figure 3)**. The Augmented Dickey-Fuller test revealed that the seasonality trend was statistically significant (P < 0.01). In 2020, likely due to the COVID-19 pandemic, no cases were recorded.

**Figure 3:**
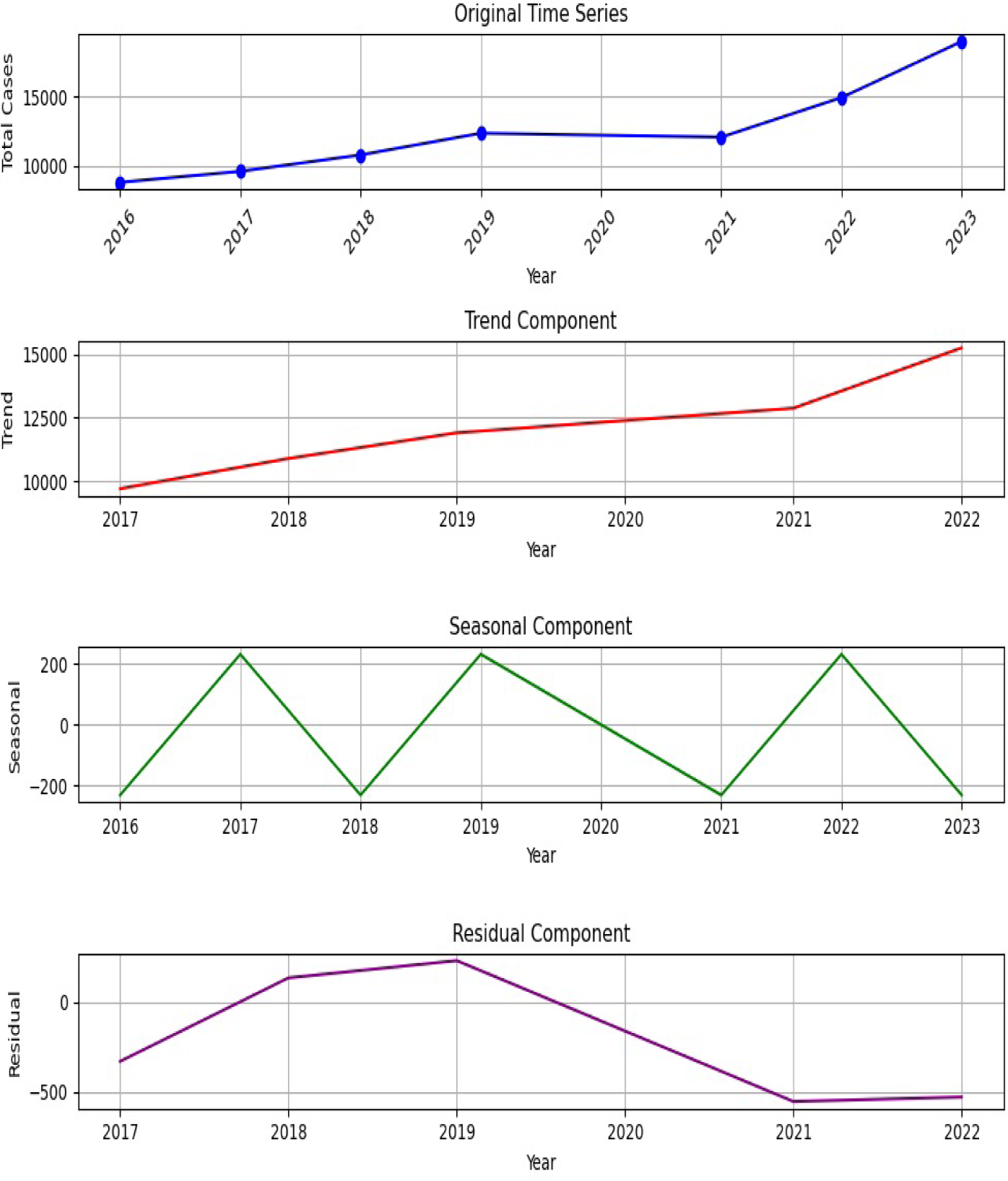
Time series analysis results showing seasonality, trend, and residuals in bites cases reported from 2016 to 2023

**Figure 4** illustrates the annual variation and increasing trend of rabid animal bite cases. The Poisson regression test indicated that the annual trend was statistically significant (P < 0.001). Notably, no cases were recorded in 2020, likely due to the impact of the COVID-19 pandemic.

**Figure 4:**
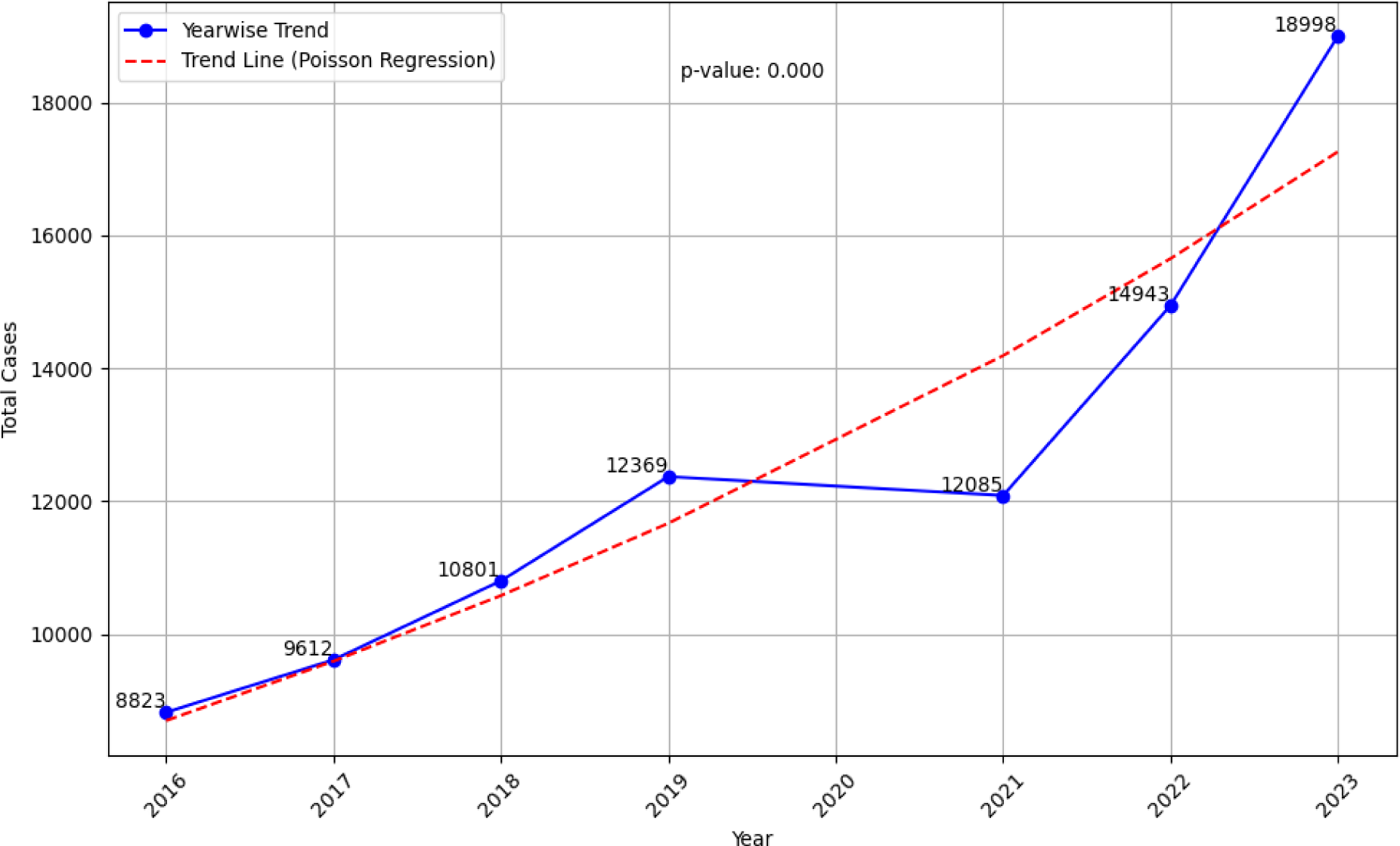
Yearly trends in rabid animal bite cases (2016–2023) reported in 2016 to 2023.

## 4. Discussion

Results of this study highlight the need for improvements in healthcare practices and treatment protocols, particularly to ensure timely vaccination delivery. The increasing trend in rabid animal bite cases highlights the need to enhance surveillance and control measures. Targeted awareness campaigns and preventive measures, specifically tailored to high-risk groups such as males, children under 10 years of age, dogs, and cats, are essential. Coordinated efforts among healthcare professionals, policymakers, and community stakeholders are critical for effectively mitigating the incidence of rabies cases, safeguarding public health, and achieving the goal of eradicating dog-mediated rabies by 2030 in the region.

The annual trends observed in this study reveal a noteworthy increase in the proportion of animal bites within the Mymensingh district over time. The rising proportion of animal bite injuries underscores the ongoing public health challenge of potential rabies exposure. These results align with several previous studies (21–23). The increasing proportion of animal bites may be attributed to rising human population and pet ownership, enhanced data recording quality, and increased community awareness of rabies prevention measures.

Given that rabies is a fatal disease, prompt administration of the vaccine after an incident is crucial. However, the majority of bite cases in this study received the vaccine within the first 24 hours after being bitten. This delay in vaccination may be attributed to a lack of awareness among the general population and a failure to recognize the importance of seeking immediate treatment following an animal bite. Post-exposure prophylaxis (PEP) stands as the most critical life-saving intervention to prevent rabies in humans following exposure (24). The average time taken to administer the vaccine after a bite is critical for the effectiveness of post-exposure prophylaxis (20, 25). Thus, efforts to increase public awareness about the urgency of seeking immediate medical attention and receiving the vaccine promptly after an animal bite are essential to enhance the timely delivery of post-exposure prophylaxis and mitigate the risk of rabies transmission.

There was a noticeable trend of incomplete vaccination courses among individuals. This may stem from limited access to post-exposure rabies vaccines at hospitals in each Upazila and lack of awareness regarding the importance of completing the full vaccine regimen. A study conducted in Vietnam highlighted a similar issue, where despite the availability of vaccines, fewer than 50% of patients completed their intramuscular vaccine regimen (26). This emphasizes the need for intensified efforts to educate the public about the significance of completing the entire vaccination course to ensure optimal protection against rabies.

This study revealed that dogs and cats were the predominant animals involved in bites, consistent with numerous findings worldwide. Dogs accounted for 76–94% of reported bite cases in low- and middle-income countries, while cats accounted for 2–50% globally (11). Additionally, the presence of monkeys, foxes, and mongoose in the study indicates a broader spectrum of animals contributing to bites. The consistency in these findings across studies underscores the reliability of the observation that dogs and cats collectively account for approximately 90% of reported animal bites. Given these findings, proactive vaccination campaigns targeting dogs and cats in high-risk zones period can significantly reduce rabies cases in the district. This approach can help mitigate the risk of rabies transmission and protect both human and animal populations from this deadly disease.

The highest proportion of bite cases was observed in the age group up to 10 years old. Children’s smaller size renders them more prone to dog and cat attacks, exacerbated by their unfamiliarity with handling pets on the street, inadvertently provoking such incidents (27). Moreover, in the home children interact with dogs and cats more commonly. This observation aligns with findings from another study, which noted that children often experience bites on their heads, necks, shoulders, and upper limbs (28). Reports from the World Health Organization (WHO) further highlight that children below the age of 15 are among the most vulnerable populations susceptible to rabies (11).

The present research findings indicate a significant gender discrepancy in animal bites, with males experiencing approximately twice as many bites as females. This observation is consistent with previous studies, which have also noted a higher incidence of animal bites among males compared to females (29, 30).

The lower extremities of the body, particularly the legs, were found to be predominant sites for animal bites. This observation is consistent with another study, which reported that the majority of bites (73.1%) occurred in the lower extremities (7).

The percentage of type 3 bites increased from 3.6% in 2022 to 6.5% in 2023. The increase may be attributed to changes in animal behavior, human population density, the popularity of domestic pets, which increase animal contact and risk, and improved reporting and monitoring systems. Human Rabies Immunoglobulin (HRIG) is a life-saving immune biological, particularly necessary for individuals exposed to category 3 animal bites. These bites are characterized by severe wounds with a high risk of rabies transmission, necessitating immediate administration of HRIG (31). HRIG provides immediate protection by supplying neutralizing antibodies. Previous studies have shown that these antibodies play a crucial role in bridging the gap until the individual’s immune system can generate vaccine-mediated antibodies (31).

The monthly distribution of rabid animal bite cases in the current study exhibited variation, with the highest number of cases reported from October to January. Earlier studies have reported similar findings of a higher percentage of rabid bite cases occurring during these months, including November, December (32). A similar trend has been observed in domestic ruminants rabies cases in Bangladesh during December, January, and July (6). This coincides with changes in animal behavior, such as breeding seasons or migration patterns, as well as increased animal activity and rabies transmission during these periods (6, 33).

The highest risk of rabid bite cases was Mymensingh Sadar, followed by Trishal. Underreporting in areas might lead to lower proportions. Rabies underreporting often occurs due to poor surveillance and diagnostic challenges, resulting in an underestimated disease burden (13). Depending exclusively on clinical diagnoses undermines the reliability of rabies surveillance systems (7). Therefore, the number of recorded animal bite cases in routine surveillance within the Mymensingh district might represent only a portion of the disease burden. Considering the challenges in diagnosing rabies in developing nations and the lack of precise human rabies trend data, surveillance data on animal bites becomes invaluable. This data offers essential insights to enhance rabies surveillance efforts and improve the allocation of medical and veterinary resources (34).

An effective rabies control strategy should prioritize enhancing collaboration and coordination across sectors using a ’One Health’ approach, as recommended during the 2015 global rabies conference. Successful control of human rabies in certain Asian regions has been attributed to robust collaboration between human and animal sectors (35). The government of Bangladesh has initiated various strategies to eliminate rabies, including advocacy, communication, and social mobilization (ACSM), modern treatment for animal bites, mass dog vaccination (MDV), and dog population management (35). However, the program’s effectiveness heavily relies on public awareness of rabies, attitudes toward animals, and informed healthcare-seeking behaviour following animal bites. Community knowledge, attitudes, and practices are vital in preventing and managing rabies, impacting human and animal populations (36).

Our study is limited by its reliance on hospital-based data collection, which might not fully capture the extent of rabies cases in the community. Hospital-based data primarily reflects individuals who seek medical care after experiencing moderate to severe animal bites, thus potentially underestimating the true prevalence of rabies within the population. Conducting community-based research would offer a more comprehensive understanding of the magnitude of the issue by including individuals who may not seek medical attention for animal bites or who may be treated outside of hospital settings. By surveying households and communities, researchers can gather data on unreported or untreated animal bites, as well as attitudes, beliefs, and practices related to rabies prevention and management.

## 5. Conclusion

The findings of our study indicate several areas where improvements are needed in healthcare practices and treatment protocols related to rabies prevention and management. One important gap identified is the delay in delivering timely vaccinations following animal bites. This delay poses a considerable risk to individuals exposed to rabies and highlights the need for simplified processes to ensure prompt vaccination delivery. The rise in rabies cases highlights the importance of strengthening disease surveillance systems to detect and report cases promptly, as well as implementing targeted interventions to prevent further transmission. To address these challenges, targeted awareness campaigns and preventive measures tailored to high-risk groups are essential. This includes educating the public, particularly males and children under 10 years old who are more susceptible to animal bites, about the importance of seeking immediate medical attention and receiving timely vaccinations after exposure to rabies. Additionally, initiatives aimed at promoting responsible pet ownership practices and increasing vaccination coverage among dogs and cats can help reduce the risk of rabies transmission within communities. Achieving these goals requires coordinated efforts among healthcare professionals, policymakers, community stakeholders and a One Health approach. Collaboration across sectors is crucial for developing and implementing comprehensive rabies control and prevention strategies. By working together, we can effectively mitigate the incidence of rabies cases, safeguard public health, and work towards the goal of eradicating dog-mediated rabies by 2030 in the region.

## Data Availability

All data generated from this study are presented within the manuscript.

## Acknowledgements

We extend our heartfelt appreciation to SKS Hospital for their invaluable assistance in furnishing the requisite data for the research. Their generosity and gracious support throughout the process are sincerely acknowledged.

## Authors’ contributions

Conceived and designed the experiments: AKMAR CSC. Data collection, input and curation: AR IS CSC. Data analysis: AR CSC AKMAR. Writing-original draft: CSC AR SB MMM. Supervision: MW MAE DPN. Writing Review & editing: MW AKMAR. All authors have read and agreed to the published version of the manuscript

## References

1. Dodds WJ, Larson LJ, Christine KL, Schultz RD. Duration of immunity after rabies vaccination in dogs: The Rabies Challenge Fund research study. Canadian Journal of Veterinary Research. 2020;84(2):153–8.

2. Mandal K, Haldar SR, Satapathy DM. Geospatial Distribution and Sociodemographic Profile of Animal Bite Cases attending Anti-rabies Clinic of a Tertiary Care Hospital in Southern Odisha. APCRI Journal. 2022;24(2):1–6.

3. Masthi N, Vairavasolai P. Estimation of animal bites using GPS and google earth in an urban low income area of Bengaluru, South India. APCRI Journal. 2016;17(II):6–10.

4. Manning SE, Rupprecht CE, Fishbein D, Hanlon CA, Lumlertdacha B, Guerra M, et al. Human rabies prevention-United states, 2008. MMWR Recomm Rep. 2008;57:1–28.

5. Amiri S, Maleki Z, Nikbakht H-A, Hassanipour S, Salehiniya H, Ghayour A-R, et al. Epidemiological Patterns of Animal Bites in the Najafabad, Center of Iran (2012–2017). Annals of Global Health. 2020;86(1).

6. Noman Z, Anika T, Haque Z, Rahman A, Ward M, Martínez-López B. Risk factors for rabid animal bites: a study in domestic ruminants in Mymensingh district, Bangladesh. Epidemiology & Infection. 2021;149:e76.

7. Rohde RE, Rupprecht CE. Update on lyssaviruses and rabies: will past progress play as prologue in the near term towards future elimination? Faculty reviews. 2020;9.

8. Pattanayak S, Malla TK, Bara BK, Behera MK. Epidemiological study of animal bite victims and admission in general surgery department, in Southern Odisha: a cross sectional institutional study. International Surgery Journal. 2017;4(10):3470–3.

9. Hossain M, Bulbul T, Ahmed K, Ahmed Z, Salimuzzaman M, Haque MS, et al. Five-year (January 2004–December 2008) surveillance on animal bite and rabies vaccine utilization in the Infectious Disease Hospital, Dhaka, Bangladesh. Vaccine. 2011;29(5):1036–40.

10. Cleaveland S, Hampson K. Rabies elimination research: juxtaposing optimism, pragmatism and realism. Proceedings of the Royal Society B: Biological Sciences. 2017;284(1869):20171880.

11. Organization WH. Zero by 30: the global strategic plan to end human deaths from dog-mediated rabies by 2030. 2018.

12. Fooks AR, Banyard AC, Horton DL, Johnson N, McElhinney LM, Jackson AC. Current status of rabies and prospects for elimination. The Lancet. 2014;384(9951):1389–99.

13. Organization WH. WHO expert consultation on rabies: second report: World Health Organization; 2013.

14. Meske M, Fanelli A, Rocha F, Awada L, Soto PC, Mapitse N, et al. Evolution of rabies in South America and inter-species dynamics (2009–2018). Tropical Medicine and Infectious Disease. 2021;6(2):98.

15. Ghosh S, Rana MS, Islam MK, Chowdhury S, Haider N, Kafi MAH, et al. Trends and clinico-epidemiological features of human rabies cases in Bangladesh 2006–2018. Scientific reports. 2020;10(1):2410.

16. Nel LH. Discrepancies in data reporting for rabies, Africa. Emerging infectious diseases. 2013;19(4):529.

17. Ghosh S, Chowdhury S, Haider N, Bhowmik RK, Rana MS, Prue Marma AS, et al. Awareness of rabies and response to dog bites in a Bangladesh community. Veterinary medicine and science. 2016;2(3):161–9.

18. Mapatse MF. Public health awareness and seroprevalence of rabies in dogs in Limpopo National Park, and the phylogeny of rabies virus in Mozambique: University of Pretoria; 2021.

19. Azad AK, Mohakhali D. National Preparedness and Response Plan for COVID-19, Bangladesh. Dhaka: Ministry of Health and Family Welfare. 2020.

20. Liu Q, Wang X, Liu B, Gong Y, Mkandawire N, Li W, et al. Improper wound treatment and delay of rabies post-exposure prophylaxis of animal bite victims in China: Prevalence and determinants. PLoS neglected tropical diseases. 2017;11(7):e0005663.

21. Fèvre EM, Kaboyo R, Persson V, Edelsten M, Coleman P, Cleaveland S. The epidemiology of animal bite injuries in Uganda and projections of the burden of rabies. Tropical medicine & international health. 2005;10(8):790–8.

22. Masiira B, Makumbi I, Matovu JK, Ario AR, Nabukenya I, Kihembo C, et al. Long term trends and spatial distribution of animal bite injuries and deaths due to human rabies infection in Uganda, 2001-2015. PloS one. 2018;13(8):e0198568.

23. Nikbakht H, Heydari H, Malakzadeh Kebria R, Yegane Kasgari M, Mirzad M, Hosseini S. Epidemiological patterns of animal bite injuries in victims under 18 year old in Babol, Iran (2010-14). Journal of Babol University of Medical Sciences. 2015;17(11):67–73.

24. Quiambao BP, Dy-Tioco HZ, Dizon RM, Crisostomo ME, Teuwen DE. Rabies post-exposure prophylaxis with purified equine rabies immunoglobulin: one-year follow-up of patients with laboratory-confirmed category III rabies exposure in the Philippines. Vaccine. 2009;27(51):7162–6.

25. Denis M, Knezevic I, Wilde H, Hemachudha T, Briggs D, Knopf L. An overview of the immunogenicity and effectiveness of current human rabies vaccines administered by intradermal route. Vaccine. 2019;37:A99–A106.

26. Tran CH, Afriyie DO, Pham TN, Otsu S, Urabe M, Dang AD, et al. Rabies post-exposure prophylaxis initiation and adherence among patients in Vietnam, 2014–2016. Vaccine. 2019;37:A54–A63.

27. Patel S, Toppo M, Lodha R. An epidemiological study of animal bite cases in a tertiary care center of Bhopal city: a cross-sectional study. Int J Med Sci Public Health. 2017;6(3):1.

28. Overall KL, Love M. Dog bites to humans—demography, epidemiology, injury, and risk. Journal of the American Veterinary Medical Association. 2001;218(12):1923–34.

29. Gebru G, Romha G, Asefa A, Hadush H, Biedemariam M. Risk factors and spatio-temporal patterns of human rabies exposure in Northwestern Tigray, Ethiopia. Annals of global health. 2019;85(1).

30. Kilic B, Unal B, Semin S, Konakci SK. An important public health problem: rabies suspected bites and post-exposure prophylaxis in a health district in Turkey. International journal of infectious diseases. 2006;10(3):248–54.

31. Haradanhalli RS, Fotedar N, Kumari N, Narayana DA. Safety and clinical efficacy of human rabies immunoglobulin in post exposure prophylaxis for category III animal exposures. Human Vaccines & Immunotherapeutics. 2022;18(5):2081024.

32. Kinge KV, Supe AC. Epidemiology of animal bite cases reported to anti-rabies vaccination OPD at a tertiary-care hospital, Nagpur. Int J Med Sci Public Health. 2016;5(8):1579–82.

33. Guo D, Yin W, Yu H, Thill J-C, Yang W, Chen F, et al. The role of socioeconomic and climatic factors in the spatio-temporal variation of human rabies in China. BMC infectious diseases. 2018;18:1–13.

34. Cleaveland S, Fevre EM, Kaare M, Coleman PG. Estimating human rabies mortality in the United Republic of Tanzania from dog bite injuries. Bulletin of the World health Organization. 2002;80(4):304–10.

35. Byrnes H, Britton A, Bhutia T. Eliminating dog-mediated rabies in Sikkim, India: a 10-year pathway to success for the SARAH program. Frontiers in veterinary science. 2017;4:28.

36. Tenzin, Dhand NK, Rai BD, Changlo, Tenzin S, Tsheten K, et al. Community-based study on knowledge, attitudes and perception of rabies in Gelephu, south-central Bhutan. International health. 2012;4(3):210–9.

